# Association of frailty, age, and biological sex with SARS-CoV-2 mRNA vaccine-induced immunity in older adults

**DOI:** 10.1101/2022.03.11.22272269

**Authors:** Janna R. Shapiro, Han-Sol Park, Tihitina Y. Aytenfisu, Christopher Caputo, John Lee, Trevor S. Johnston, Huifen Li, Pricila Hauk, Henning Jacobsen, Yukang Li, Engle Abrams, Andrew J. Kocot, Tianrui Yang, Yushu Huang, Steven M. Cramer, Michael J. Betenbaugh, Amanda K. Debes, Rosemary Morgan, Aaron M. Milstone, Andrew H. Karaba, Sean X. Leng, Sabra L. Klein

## Abstract

**Background:** Male sex and old age are risk factors for severe COVID-19, but the intersection of sex and aging on antibody responses to SARS-CoV-2 vaccines has not been characterized.

**Methods:** Plasma samples were collected from older adults (75-98 years) before and after three doses of SARS-CoV-2 mRNA vaccination, and from younger adults (18-74 years) post-dose two, for comparison. Antibody binding to SARS-CoV-2 antigens (spike protein [S], S-receptor binding domain [S-RBD], and nucleocapsid [N]) and functional activity against S were measured against the vaccine virus and variants of concern (VOC).

**Results:** Vaccination induced greater antibody titers in older females than males, with both age and frailty associated with reduced antibody responses to vaccine antigens in males, but not females. ACE2 binding inhibition declined more than anti-S or anti-S-RBD IgG in the six months following the second dose (28-fold vs. 12- and 11-fold decreases in titer). The third dose restored functional antibody responses and eliminated disparities caused by sex, age, and frailty in older adults. Responses to the VOC were significantly reduced relative to the vaccine virus, with older males having lower titers to the VOC than females. Older adults had lower responses to the vaccine and VOC viruses than younger adults, with disparities being greater in males than females.

**Conclusion:** Older and frail males may be more vulnerable to breakthrough infections due to low antibody responses before receipt of a third vaccine dose. Promoting third dose coverage in older adults, especially males, is crucial to protecting this vulnerable population.

**Brief summary:** SARS-CoV-2 mRNA vaccination induces greater antibody response in older females than males, and age and frailty reduce responses in males only. These effects are eliminated by a third vaccine dose, highlighting the need for third dose coverage, especially in older males.

## Introduction

The disproportionate burden of COVID-19 in older adults was recognized early in the pandemic [1-3]. The phase III trials for the two mRNA vaccines (mRNA-1273 and BNT162b2) revealed high efficacy in older adults [4, 5], for whom immunosenescence is thought to impair vaccine-induced immune responses [6]. Clinical trials, however, often fail to represent the oldest and frailest subset of the population. Accordingly, wide-spread use of the vaccine in long-term care facility residents revealed that old age is a risk factor for poor antibody responses [7-9].

Male sex also is a significant predictor of severe COVID-19 outcomes at older ages [10-14]. There is extensive evidence that the effects of aging on the immune system differ between the sexes, including that immunosenescence occurs at a slower rate in females than males [15, 16]. The implications of biological sex are evident in the response to repeated seasonal influenza vaccination in older adults, where pre-vaccination titers decrease with age in males but not in females, suggesting that older females enter each influenza season with greater immunity than their male counterparts [17]. In the context of SARS-CoV-2 vaccines, however, studies have failed to provide sex-disaggregated data within each age group [18, 19], and little is known about how biological sex may modify the effects of age, and age-related factors like frailty, on vaccine immunogenicity. Here, we investigate sex differences and sex-specific effects of aging in the humoral immune response to the vaccine virus and variants of concern (VOC) induced by three doses of a SARS-CoV-2 mRNA vaccine in a cohort of adults above 75 years of age. We illustrate that the age- and frailty-associated declines in antibody responses occur to a greater extent in males than females.

## Methods

### Cohorts

Older adults (75-98 years) were recruited from the Johns Hopkins Longitudinal Influenza Immunization Study of Aging over 75 years of age (JH LIISA 75+) cohort [17] (**Table 1**). Individuals who had worsening or new-onset of immune-modulating conditions (e.g., rheumatoid arthritis, hematologic malignancies, or other cancers) or a previous diagnosis of COVID-19 were excluded. Participants came to the Johns Hopkins Bayview Medcal Center, or study visits were conducted at participants’ homes, as needed. At pre-vaccination visits (Pre), frailty status was assessed using the Fried Frailty Phenotype [20] and a baseline blood draw was obtained. Subsequent receipt of two (primary vaccination series) or three doses of a SARS-CoV-2 mRNA vaccine, either mRNA-1273 or BNT162b2, was confirmed via vaccination cards, and blood samples were collected 14-30 days (i.e., average ≤ 1 month [M]) post dose 1 (<1M_PD1)), 14-30 days post dose 2 (<1M_PD2), 90 (± 15) days post dose 2 (3M_PD2), 180 (± 15) days post dose 2 (6M_PD2), and 14-60 days post dose 3 (1M_PD3).

**Table 1.**
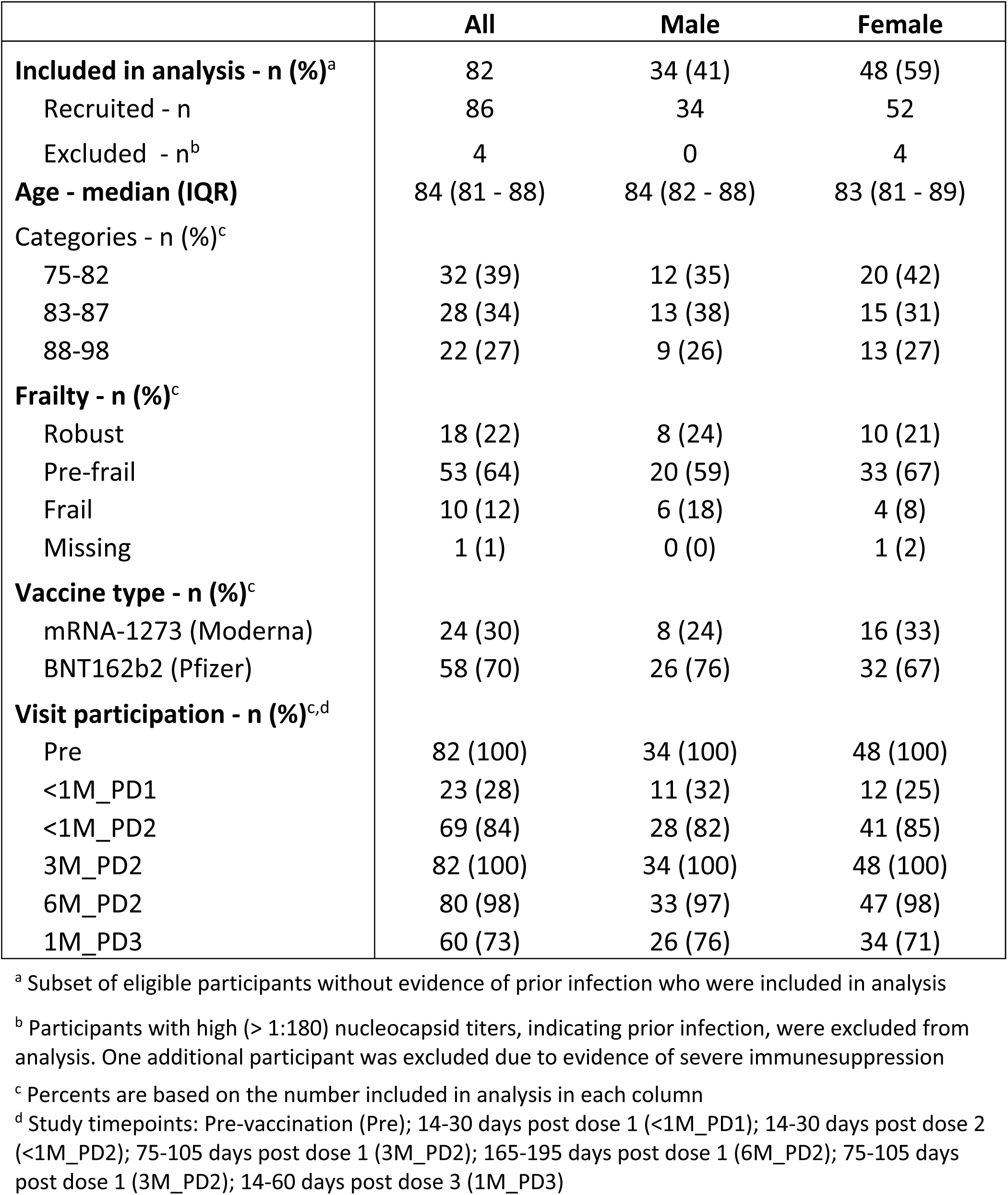
Older adult participant characteristics.

Younger adult healthcare workers from the Johns Hopkins Health System were also sampled as a comparison group. Recruitment of these younger adults has been reported elsewhere [21]. To be eligible for the present study, participants needed to be younger than 75 years, not have a history of COVID-19, and have two samples collected at least 90 days apart, with the first one collected at least 14 days after receiving the second dose of a SARS-CoV-2 mRNA vaccine. Due to low plasma volumes, these amples were not tested for ACE2-inhibition, and data are missing for antibody titration against antigens from VOC for some participants. Exact sample sizes are included in figure legends. For both cohorts, written, informed consent was obtained from all participants, and the study protocols were approved by the Johns Hopkins School of Medicine Institutional Review Board.

### Laboratory methods

Detailed ELISA and ACE2 inhibition methods can be found in the **Supplemental materials**. Briefly, plasmids expressing recombinant nucleocapsid (N), Spike (S), or S receptor-binding domain (S-RBD) of the vaccine strain and the Alpha, Delta, and Omicron variants of SARS-CoV-2 were engineered at Johns Hopkins as described previously [22] or obtained through the NCI Serological Sciences Network for COVID-19 [23] (**Supplemental Table 1**). Recombinant proteins were used to coat plates for indirect ELISA measuring plasma IgG against N, S, or S-RBD. Results were expressed as the log10-transformed area under the curve (AUC) generated from ten three-fold serial plasma dilutions, as previously described [22]. The ability of plasma antibodies to inhibit ACE2 binding to S was measured using Meso Scale Diagnostics (MSD) V-PLEX SARS-CoV-2 ACE2 kits according to the manufacturer’s protocol at a dilution of 1:100 [24]. Data were expressed as the log10-transformed concentration (μg/ml) of ACE2-inhibiting antibodies (ACE2iAb), which are equivalent to anti-S monoclonal antibodies. IgG binding to seasonal and epidemic β-coronavirus S proteins were measured using the multiplex chemiluminescent MSD V-PLEX COVID-19 Coronavirus Panel 3 (IgG) Kit according to the manufacturer’s protocol at a dilution of 1:5000.

### Statistical methods

Longitudinal data in the older adult cohort were analyzed using mixed-effects models with random intercepts on the individual to account for repeated measures, and interaction terms between study timepoint (categorical) and sex (self-report), age (categorized based on terciles) and frailty status. Linear regression models including interaction terms between sex and age or frailty were used to investigate sex-specific effects at individual timepoints. To compare the older and younger cohorts, the number of days post-dose 2 was used as a continuous predictor and cubic splines were included to study non-linear relationships [25]. Cubic spline knots were placed at 30-, 100-, and 160-days post-vaccination, points chosen to approximately divide the data into quartiles. Mixed-effects models included an interaction term between time and cohort and were repeated separately for males and females. Differences between cohorts were tested at three sentinel points (14-, 90- and 180-days post dose 2). All p-values <0.05 were considered statistically significant. Analyses were performed in Stata 15 (StataCorp).

## Results

### Study population demographics

Eighty-six older adults were recruited from the Baltimore area, with three participants excluded from analysis due to high N titers (i.e., titer >180), suggesting prior infection (**Supplemental figure 1**). One additional participant was excluded from analysis due to evidence of severe immunosuppression, such that their responses could not be accurately captured in population-level models. Characteristics of the 82 participants included in analysis are detailed in **Table 1**. The population had more females (59%) than males, and a median age of 84 years. Most participants were classified as pre-frail (64%) and a greater percentage of males than females were frail. All participants received two doses of a SARS-CoV-2 mRNA vaccine, with the majority (70%) receiving BNT162b2. Sixty participants (73%) received a third vaccine dose at least six months after the second dose.

Demographic information for the younger adult cohort is provided in **Table 2**. Of 84 eligible participants from the affiliated study [21], three were excluded due to high anti-N titers (**Supplemental figure 1**). In the younger population included in analysis, there were more females than males (60% vs 40%), most participants were between 30 and 49 years of age, and a majority of samples were collected 21-43 days and 125-150 days after receipt of the second vaccine dose.

**Table 2.** Younger adult participant characteristics.

|  | **All** | **Male** | **Female** |
| --- | --- | --- | --- |
| **Included in analysis - n (%)<sup>a</sup>** | 81 | 32 (40) | 49 (60) |
| Eligible - n<sup>b</sup> | 84 | 32 | 52 |
| Excluded - n<sup>c</sup> | 3 | 0 | 3 |
| **Age at vaccination - n (%)<sup>d</sup>** |  |  |  |
| ≤29 | 14 (17) | 4 (12) | 10 (20) |
| 30-39 | 32 (40) | 11 (34) | 21 (43) |
| 40-49 | 18 (22) | 8 (25) | 10 (20) |
| 50-59 | 7 (9) | 4 (13) | 3 (6) |
| 60-74 | 10 (12) | 5 (16) | 5 (10) |
| **Sample 1 - days post dose 2** |  |  |  |
| Mean (min-max) | 33 (16-76) | 31 (16-65) | 34 (16-76) |
| Median (IQR) | 29 (21-43) | 27 (21-41) | 29 (21-43) |
| **Sample 2 - days post dose 2** |  |  |  |
| Mean (min-max) | 138 (96-190) | 142 (110 - 190) | 136 (96-183) |
| Median (IQR) | 137 (125-150) | 139 (128-156) | 137 (123-147) |

<sup>a</sup> Subset of eligible participants without evidence of prior infection who were included in analysis
<sup>b</sup> Eligible participants from affiliated study (ref) were <75 years of age, had remaining serum from 2 samples collected at least 90 days apart 14-200 days following 2 doses of an mRNA SARS-CoV-2 vaccine who did not report prior SARS-CoV-2 infection
<sup>c</sup> Participants with high (> 1:180) nucleocapsid titers, indicating prior infection, were excluded from analysis
<sup>d</sup> Percents are based on the number included in analysis in each column

### Older females mount greater responses to vaccination than older males

Among older adults, IgG binding to S and S-RBD of the vaccine strain increased significantly in response to the first two vaccine doses and then decreased significantly in the 6 months following immunization (p<0.001 for all comparisons; **Figure 1A-B & D**). Geometric mean titers (GMT) decreased 11- and 12-fold, for S and S-RBD, respectively, from <1M_PD2 to 6M_PD2 (**Supplemental Table 2**). Females mounted greater IgG responses to S and S-RBD relative to their baseline than males at all post-vaccination timepoints (p<0.02 for all comparisons, **Figure 1A-B & D**). Older females also had greater titers of IgG against S and S-RBD at each visit, and this difference was significant for anti-S IgG at <1M_PD1 (p=0.020) and at 3M_PD2 (p=0.026). After receipt of a third vaccine dose, anti-S and S-RBD titers increased significantly in both males and females (p<0.001), leading to GMT that were 2- and 4-fold greater than the post-dose 2 peak for S and S-RBD, respectively (**Supplemental Table 2**).

**Figure 1.**
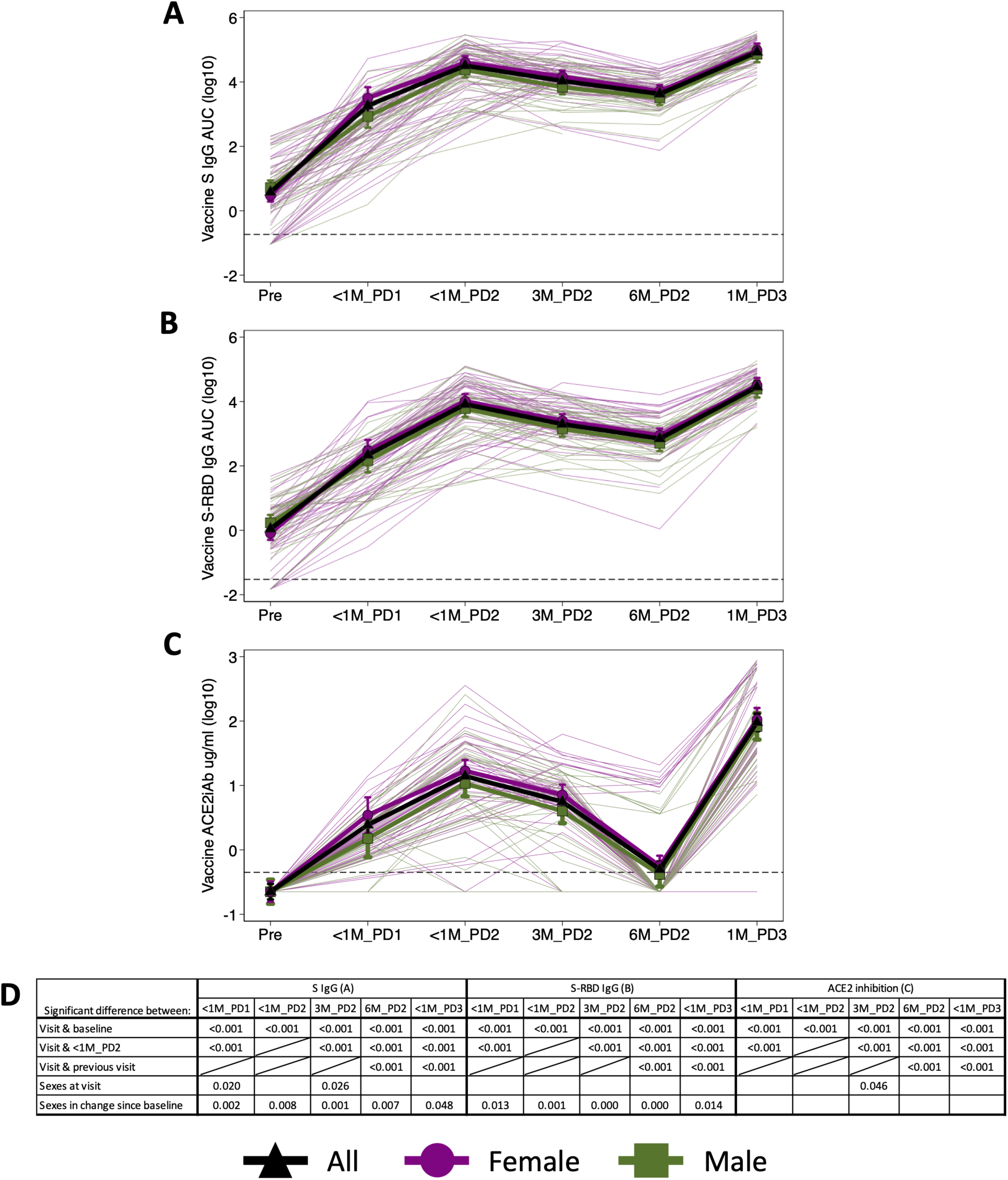
Older females mount greater humoral responses to SARS-CoV-2 mRNA vaccines than older males. Anti-spike (S) IgG (**A**), S receptor-binding domain (S-RBD) IgG (**B**), and ACE2-inhibiting antibodies (ACE2iAb) (**C**) against the vaccine strain of SARS-CoV-2 were measured at six timepoints: pre-vaccination (n= 82: 48 females, n = 34 males), 14-30 days post dose 1 (<1M_PD1; n=23: 12 females, 11 males), 14-30 days post dose 2 (<1M_PD2; n=69: 41 females, 28 males), 3 months post dose 2 (3M_PD2; n=82: 48 females, 34 males), 6 months post dose 2 (6M_PD2; n=80: 47 females, 33 males), and 14-30 days post dose 3 (1M_PD1; n=60: 34 females, 26 males). Differences between timepoints were tested using mixed-effects models with study timepoint as a dummy variable and random intercepts on the individual. Sex differences were tested using an expanded mixed-effects model that included a main effect for sex and an interaction term between sex and study timepoint. All point estimates are shown with error bars indicating the 95% confidence interval. All p-values <0.05 are reported in **D**, where blank cells indicate a p-value >0.05 and crossed out cells indicate that the comparison is reported elsewhere in the table. Dashed lines show the limits of detection.

The functional ability of antibodies to inhibit S from binding to ACE2 followed similar kinetics as anti-S IgG in response to the primary immunization series, but then decreased more rapidly in the 6-months following immunization, resulting in a 28-fold decrease in GMT from <1M_PD2 to 6M_PD2 (**Figure 1C & Supplemental Table 2**). By 6M_PD2, 79% of males and 77% of females had undetectable ACE2iAb. Sex differences were apparent at all timepoints and were significant at 3M_PD2, with females mounting stronger responses than males (p=0.046). Post dose 3, all but one participant had detectable ACE2iAb, and the geometric mean was 7-fold higher than the post-dose 2 peak (**Supplemental Table 2**). Despite differences in kinetics over time between the binding and functional assays, the three readouts of humoral immunity correlated well with each other (**Supplemental figure 2**). As expected, correlations became weaker at the lower range of the ACE2-inhibition assay. Taken together, these data suggest that older females mount stronger response to SARS-CoV-2 vaccination than males, and that a third vaccine dose is necessary to boost functional antibody responses in this population.

### The effects of age and frailty are greater in males than in females

We next assessed the overall and sex-specific effects of age on the humoral response to vaccination. Among all older participants, age was significantly associated with reduced anti-S IgG, anti-S-RBD IgG, and ACE2iAb in the six months following the primary vaccination series (**Figure 2A-C**). This effect was largely driven by the oldest tercile of the population (≥88 years). The percent of participants with undetectable ACE2iAb by 6M_PD2 increased from 67% in the youngest tercile (75-82 years) to 85% in the oldest tercile. In sex-disaggregated analyses focusing on the 3M_PD2 timepoint (i.e., a time point when all study participants were represented), age significantly impaired responses in males, but not females, leading to statistically significant sex differences in the effect of age for anti-S IgG (p=0.025) and ACE2iAb (p = 0.001; **Figure 2D-F**). The trend of greater age effects in males than females was consistent at other timepoints following the primary immunization series (**Supplemental figure 3A-F**), and by 6M_PD2, 100% of males in the oldest age group, compared to 77% of females, had undetectable ACE2iAB. After receipt of a third dose, the effect of age was no longer significant in the overall population or within either sex, suggesting that a third vaccine dose eliminated sex and age disparities in vaccine-induced immunity (**Figure 2A-C & Supplemental figure 3G-I**).

**Figure 2.**
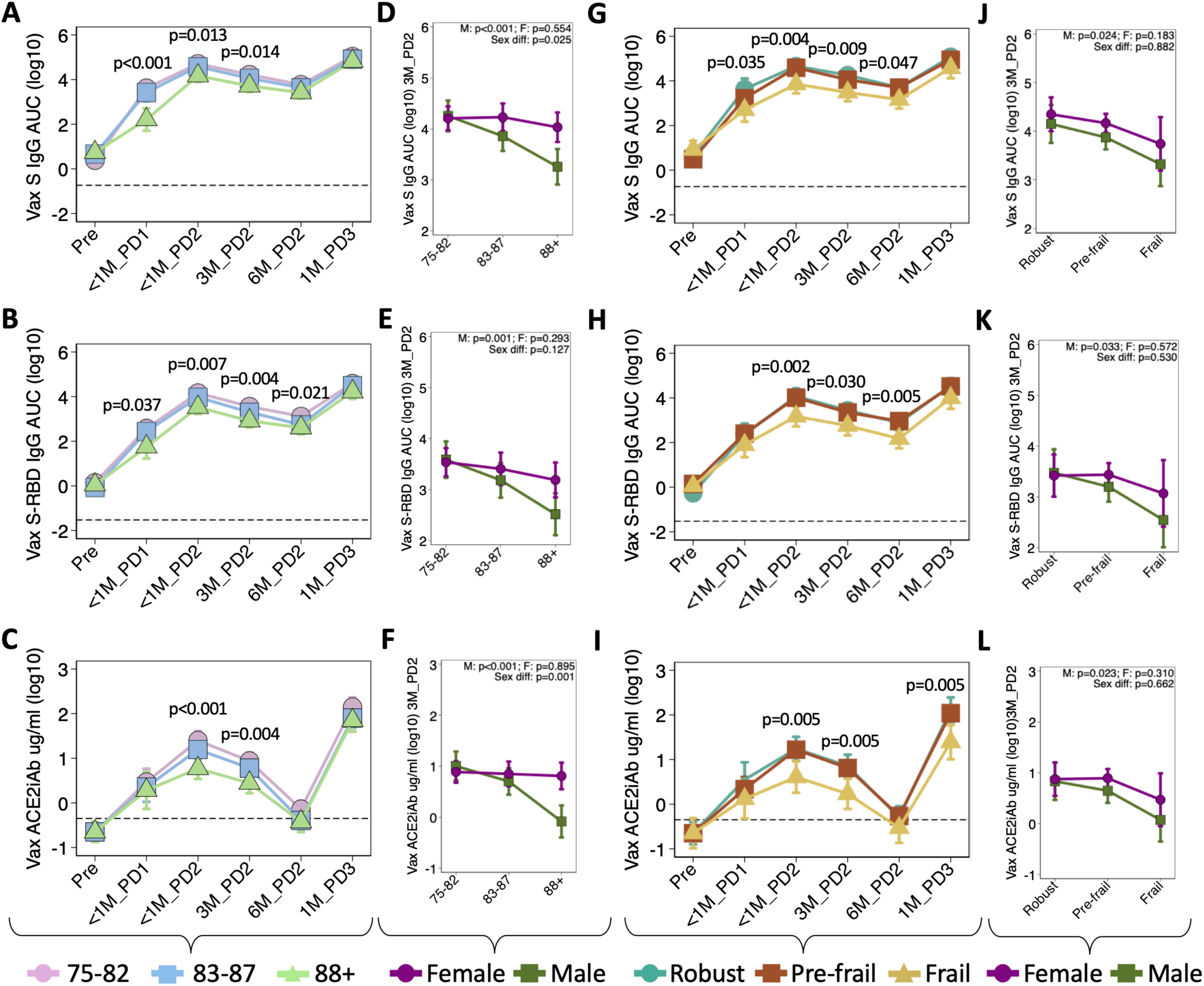
Age and frailty impact the antibody response to SARS-CoV-2 mRNA vaccines in a sex-specific manner among older adults. The effect of age on antibody kinetics is shown for Anti-spike (S) IgG (**A**), S receptor-binding domain (S-RBD) IgG (**B**), and ACE2-inhibiting antibodies (ACE2iAb) (**C**) against the vaccine strain of SARS-CoV-2. Data are shown for six timepoints: pre-vaccination (n=32 aged 75-82; n=28 aged 83-87; n=22 aged ≥88), <1M_PD1 (n=10 aged 75-82; n=8 aged 83-87; n = 5 aged ≥88), <1M_PD2 (n=24 aged 75-82; n=25 aged 83-87; n=20 aged ≥88), 3M_DP2 (n=32 aged 75-82; n=28 aged 83-87; n=22 aged ≥88), 6M_PD2 (n=31 aged 75-82; n=28 aged 83-87; n=21 aged ≥88), and 1M_PD3 (n=22 aged 75-82; n=21 aged 83-87; n=17 aged ≥88) (**A-C**). Sex-specific effects of age at 3M_PD2 are shown separately for females (n=20 aged 75-82; n=15 aged 83-87; n=13 aged ≥88) and males (n=12 aged 75-82; n=13 aged 83-87; n=9 aged ≥88) (**D-F**). The effect of frailty on antibody kinetics is shown for the three assays at six timepoints: pre-vaccination (n=18 robust; n=53 pre-frail; n=10 frail), <1M_PD1 (n=6 robust; n=12 pre-frail; n=5 frail), <1M_PD2 (n=15 robust; n=45 pre-frail; n=9 frail), 3M_PD2 (n=18 robust; n=53 pre-frail; n=10 frail), 6M_PD2 (n=18 robust; n=52 pre-frail; n=10 frail), and 1M_PD3 (n=14 robust; n=39 pre-frail; n=7 frail) (**G-I**). are shown separately for females (n=10 robust; n=33 pre-frail; n=4 frail) and males (n=8 robust; n=20 pre-frail; n=6 frail) (**J-L**). The overall effects of age (**A-C**) or frailty (**H-I**) at each timepoint were tested using mixed-effects models including a main effect for age/frailty, an interaction term between age/frailty and study timepoint, and all p-values <0.05 are shown. At 3M_PD2, the effect of age (**D-F**) or frailty (**J-L**) in males and females, and sex-differences in these effects, were tested using linear regression models with interaction terms between sex and age or frailty, and all p-values are shown. Point estimates are shown with 95% confidence intervals, and dashed lines indicate the limit of detection.

Frailty had an important overall effect, with frail participants mounting significantly weaker responses to vaccination than robust and pre-frail participants (**Figure 2G-I**). By 6M_PD2, 90% of frail participants had undetectable ACE2iAb, compared to 75% of pre-frail and robust participants. Like with age, the effect of frailty at 3M_PD2 was significant in males, but not females for all assays (**Figure 2J-L**). No significant sex differences in the effect of frailty were observed, however, and trends were less consistent over time (**Supplemental figure 3J-R**). The effect of frailty was also attenuated by the third dose but remained significant for ACE2iAb (p=0.005; **Figure 2I**). From these data, we conclude that the effects of age and frailty in older adults are largely driven by males, not females.

### Antibody responses to VOC are reduced relative to the vaccine virus

The breadth of vaccine-induced immunity in older adults was assessed by measuring anti-S IgG to the Alpha, Delta, and Omicron variants. Antibody titers to the Alpha and Delta variants were similar to each other and were both significantly reduced relative to the vaccine virus (2-4-fold lower GMT, p<0.001; **Figure 3A & Supplemental Table 3**). Titers to Omicron were further reduced relative to the vaccine virus (>5-fold difference in GMT) and the Alpha and Delta variants (p<0.001 for all comparisons; **Figure 3A & Supplemental Table 3**). Differences between anti-S IgG to the vaccine virus and the VOC were attenuated at 1M_PD3 (fold difference in GMT <1.5 for Alpha and Delta and <4 for Omicron) but remained significant (p<0.0001 for all comparisons). At 1M_PD3, anti-Alpha S IgG was significantly higher than anti-Delta S IgG (p<0.001). ACE2iAb to the Alpha and Delta variants also tended to be lower than to the vaccine strain, but these differences were smaller in magnitude than for anti-S IgG and were not statistically significant at all timepoints (**Figure 3B-C**). In sex-disaggregated analyses, females had higher responses to the VOC than males, and this difference was significant for anti-Delta S IgG (p=0.038) and ACE2iAb against the Alpha variant (p=0.034) at 3M_PD2 (**Figure 3D-E & Supplemental figure 4**).

**Figure 3.**
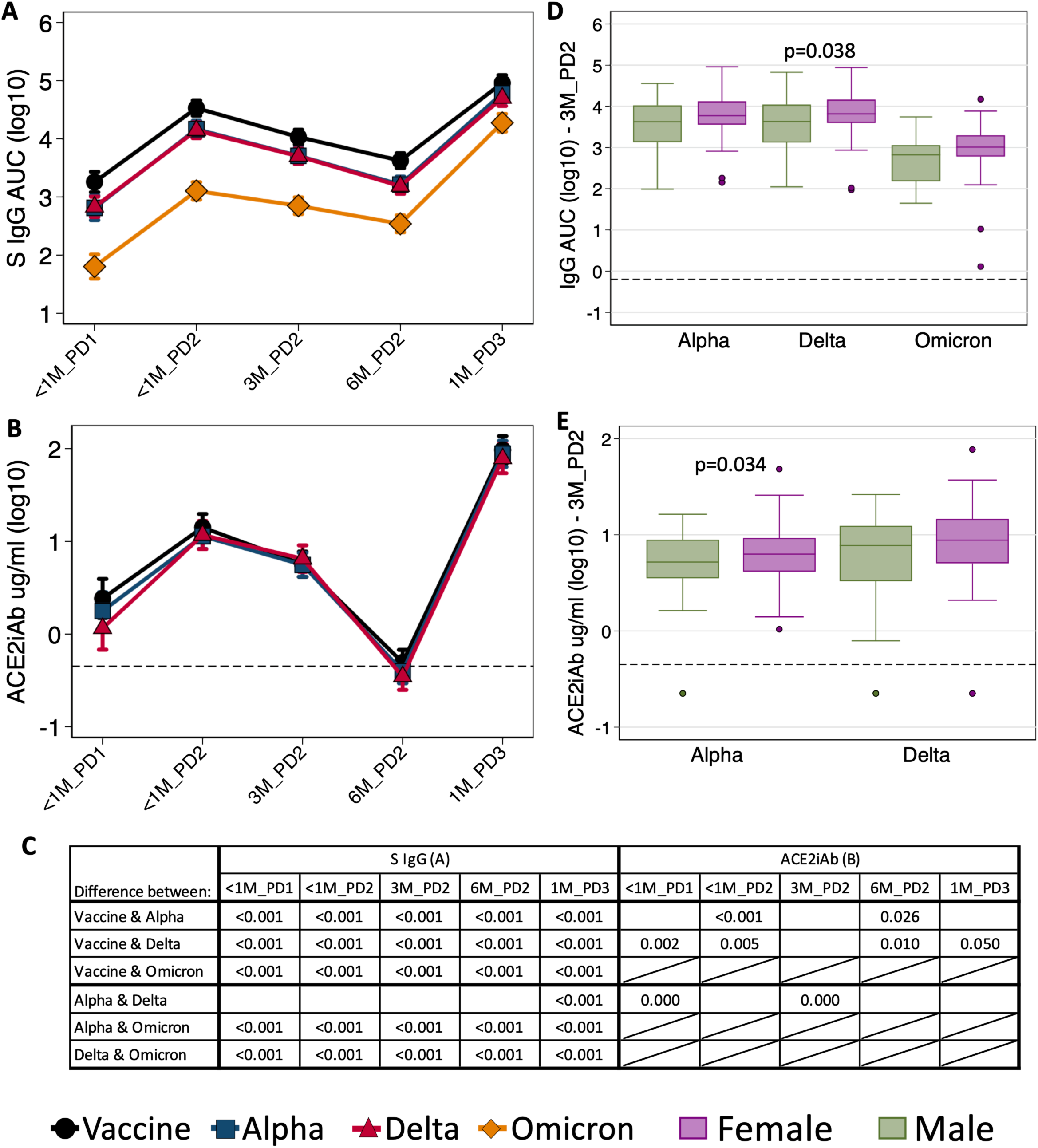
Antibody responses to Alpha, Delta, and Omicron variants are reduced relative to the vaccine virus in older adults. Anti-S (**A**), and ACE2-inhibiting (**B**) antibodies against the vaccine, Alpha and Delta strains of SARS-CoV-2 were measured post-vaccination, with symbols indicating point estimates and error bars indicating the 95% confidence interval. Differences in the responses between viral strains were measured using paired t-tests, and all p-values <0.05 are shown in **C**, where empty cells indicate p-value>0.05 and crossed-out cells indicate that the comparison was not tested. Sex-disaggregated data from the 3-month timepoint are shown, and significant sex differences are indicated by p-values (**D-E**).

### SARS-CoV-2 vaccination induces IgG responses to other β-coronaviruses

To investigate the crosss-reactivity of the vaccine-induced humoral response, IgG titers to seasonal and epidemic β-coronaviruses were measured in the older adults. Anti-HKU1 IgG titers increased in response to the primary vaccination series, but fell by 3M_PD2 (**Figure4A**). Titers of IgG recognizing OC43, MERS-CoV, and SARS-CoV-1 increased significantly in plasma samples collected after SARS-CoV-2 vaccination and remained elevated above baseline levels for 6 months (**Figure 4B-D**). Furthermore, titers to all four β-coronaviruses were significantly elevated by the third vaccine dose (**Figure 4A-D**). Notably, IgG binding to the two seasonal β-coronaviruses (HKU1 and OC43) was elevated at baseline, indicating widespread exposure to these viruses in this population. Increased antibody titers against related β-coronaviruses following SARS-CoV-2 vaccination suggests possible “back-boosting”, as observed for influenza vaccines [26] and in response to SARS-CoV-2 vaccination in the general adult population [27, 28]. These results indicate that SARS-CoV-2 vaccination increases broad antibody responses to diverse β-coronaviruses in older adults.

**Figure 4.**
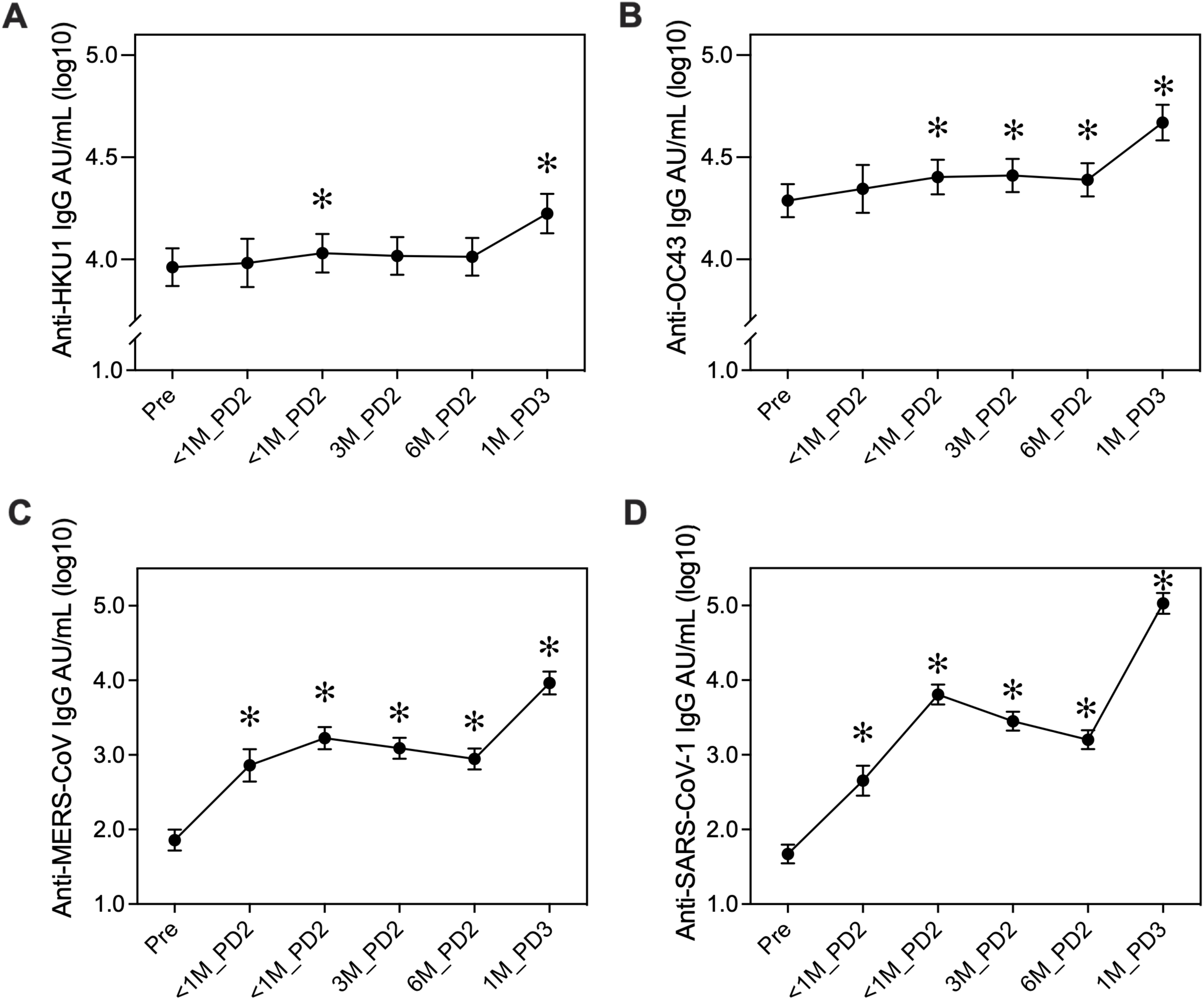
Antibody responses against seasonal and pandemic β-coronaviruses are boosted by SARS-CoV-2 vaccination in older adults. IgG specific to the spike proteins of the HKU1 (**A**), OC43 (**B**), MERS-CoV (**C**), and SARS-CoV-1 (**D**) were measured before and at five timepoints after receipt of a SARS-CoV-2 mRNA vaccine. Differences between timepoints were tested using multi-level models with study timepoint as a dummy variable and random intercepts on the individual to account for repeat measures. All point estimates are shown with error bars indicating the 95% confidence interval and asterisks indicate significant (p<0.05) increases relative to the pre-vaccination vaccination timepoint.

### Differences between older and younger cohorts are driven by males

To further investigate the sex-specific effects of aging, antibody kinetics against vaccine, Alpha, and Delta antigens were compared between the younger and older adult cohorts in the six months following the primary vaccination series. In the whole population, anti-vaccine S IgG was significantly lower in older than younger adults (p<0.001 at 14 days post-vaccination, p=0.004 at 90 days, and p=0.026 at 180 days) (**Figure 5A**). In sex-disaggregated analyses, differences between the older and younger adults were significant among males at all three sentinel points (p=0.004 at 14 days post-vaccination, p=0.005 at 90 days, and p=0.019 at 180 days), but only significant among females at 14-days post vaccination (p=0.004) (**Figure 5B-D**). In addition, the magnitude of the difference between the mean of the older cohort and the mean of younger cohort was consistently larger for males than for females across the three sentinel points (**Figure 5D**). Similar results were observed for anti-Alpha and Delta S IgG (**Figure 5E-L**). There were no significant differences in the rate of waning between older and younger adults, suggesting that antibody kinetics are not age-dependent.

**Figure 5.**
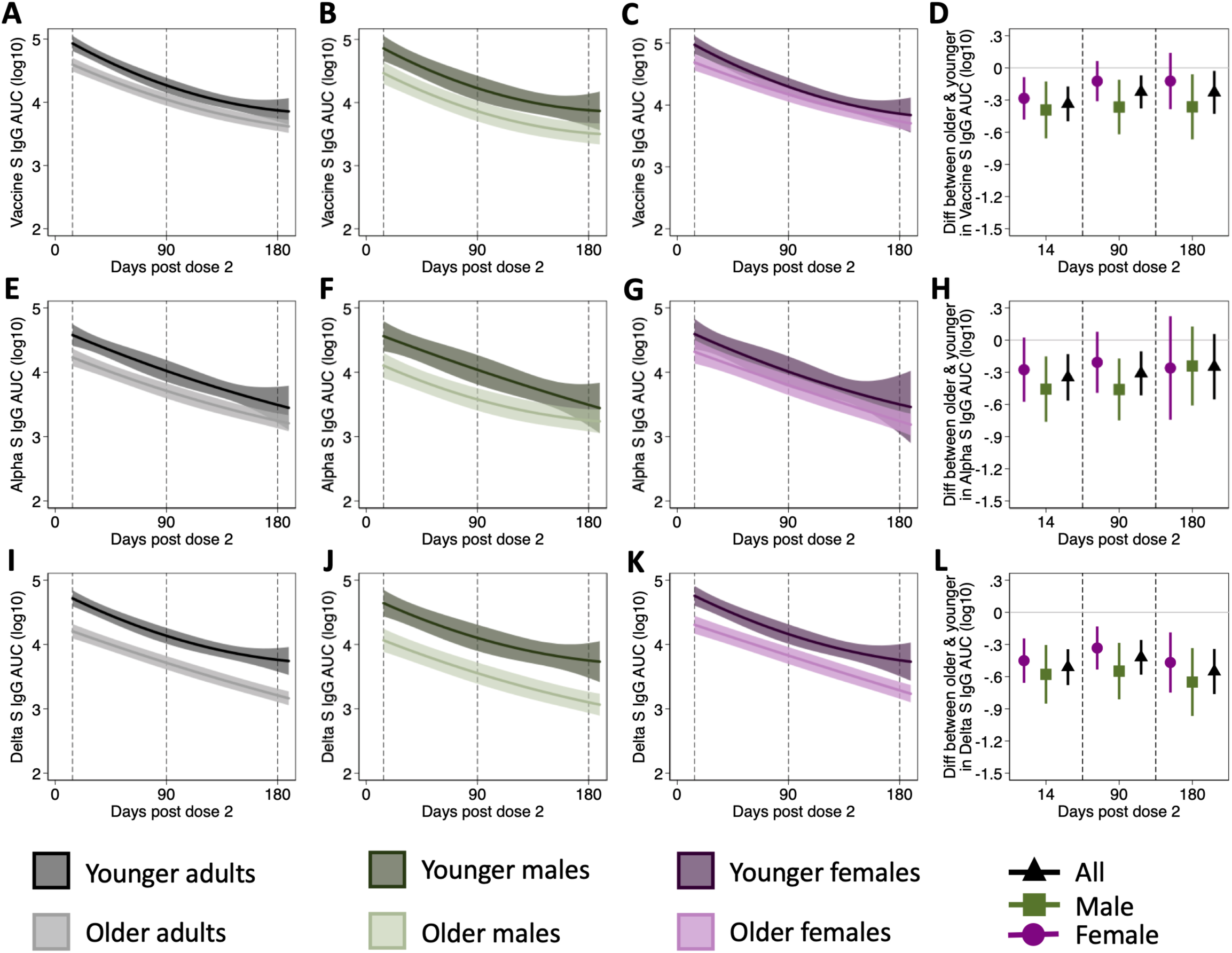
Differences between younger and older adults in antibody responses to the vaccine strain of SARS-CoV-2 are sex-dependent. Plasma samples were collected from older adults at 3 timepoints after the primary vaccination series and from younger adults at 2 post-vaccination timepoints: 16-76 days post dose 2 (early) and 96-190 days post dose 2 (late). Differences in anti-vaccine strain S IgG levels over time were compared between all younger and older adults (**A**), males (younger: n=27 early; n=30 late) (**B**), and females (younger: n=48 early; n=48 late) (**C**), and summarized at three sentinel points (14-, 90- and 180-days post-vaccination) (**D**). Comparisons of the anti-Alpha S IgG response between the younger and older groups are shown for the whole population (**E**), males (younger: n=27 early; n=26 late) (**F**), and females (younger: n=39 early; n=34 late) (**G**), with differences summarized at 14-, 90- and 180-days post-dose 2 (**H**). Comparisons of the anti-Delta S IgG response between the younger and older groups are shown for the whole population (**I**), males (younger: n=27 early; n=26 late) (**J**), and females (younger: n=47 early; n=42 late) (**K**), with differences at summarized 14-, 90- and 180-days post-dose 2 (**L**). Kinetics were analyzed using mixed-effects models with fixed effects including days post-dose 2 as a continuous predictor with cubic B-splines (knots at 30-, 100-, and 160-days post-vaccination). Shaded areas indicate 95% confidence intervals (**A-C, E-G, I-K**). In **D, H**, and **L**, point estimates for the difference between cohorts are shown with 95% confidence intervals, such that confidence intervals that do not span the null value of zero are statistically significant.

## Discussion

In this longitudinal study, older adult females mounted stronger antibody responses to SARS-CoV-2 mRNA vaccination than older males, and age and frailty were associated with reduced responses in males but not females. While the kinetics of antibody waning in the six months following immunization were not age-dependent, older adults mounted weaker initial responses to vaccination, such that their antibody titers remained lower than younger adults throughout the follow-up period. A sex-specific effect of age was observed both within the older cohort and when comparing younger and older adults, in which age-associated reductions in humoral immunity were greater among males than females. In the older adult cohort, receipt of a third vaccine dose largely eliminated disparities caused by sex, age, and frailty in antibody responses, with the exception of ACE2iAb, which remained lower in frail compared to non-frail or pre-frail participants. The effect of age on SARS-CoV-2 vaccine responses has been studied [7-9, 29-33], but the sex differential impact of age has not been reported previously. Furthermore, studies investigating frailty have not found an effect on antibody responses [34-36], but have reported that frailty increases the risk of post-vaccination breakthrough infection [37, 38], suggesting that the immunogenicity studies may have been under-powered to observe an effect of frailty or that lack of consideration of biological sex obscured the effect.

The inclusion of three measures of humoral immunity and four SARS-CoV-2 viruses allowed us to capture the breadth and depth of vaccine responses in this vulnerable population. For measurement of functional antibodies, ACE2-inhibition assays are a strong surrogate of live virus neutralization [24, 39]. In terms of responses to VOC, the reductions in anti-S IgG to the Alpha and Delta variants observed in the older adults were similar to other reports in the general population [40]. For the Omicron variant, while reductions in live-virus neutralization in vaccinated serum from the general adult population have been reported, there were no reductions in anti-Omicron S IgG [41, 42]. Given the importance of neutralizing and non-neutralizing functions of IgG in conferring protection against SARS-CoV-2 [43, 44], the markedly lower anti-Omicron S IgG level in older adults, that persisted after receipt of a third vaccine dose, suggests that this population may be more vulnerable to disease caused by the Omicron variant than younger adults, and that reformulation of vaccines to target the Omicron variant would be beneficial.

Our study had several strengths and limitations. Some of the sex-specific effects observed were differences among males that were absent among females, without statistical evidence of a sex difference (i.e., non-significant sex interaction terms) [45]. It is important to note that our findings were generated from *post-hoc* analyses that were not necessarily powered to investigate sex differences, and conclusions are limited by small samples sizes in certain sub-groups. Given the small sample sizes, it is not surprising that statistically significant sex differences were not consistently observed. Particularly for age-based analyses, however, the consistency of trends between assays and timepoints, coupled with statistically significant sex differences in the effect of aging at 3M_PD2, lend credibility to the conclusion that the effects of age on antiviral antibody responses are driven by males. Further supporting these findings are similar sex-specific effects of age observed following seasonal influenza vaccination in both younger and older adults [17, 46]. While it is important to not over-interpret ‘within-sex’ differences as ‘between-sex’ differences [47], there is considerable value in studying differences within males or feamles [48, 49]. This is particularly true given the uniqueness of the community-dwelling older adult cohort, which represent the ‘oldest’ old subset, and are distinct from the population of long-term care facility residents that has been the focus of much of the SARS-CoV-2 research in older adults.

There also were missing data in the older adult cohort, particularly at the <1M_PD1 timepoint. These missing data did not, however, depart from the missing at random assumption, and thus multi-level models were used to account for missingness. Second, the timing of sample collection was different in the older and younger cohorts. To account for this, analyses that compared the two groups used days post-vaccination as a continuous variable. Finally, we have not investigated the functional antibody responses to the Omicron variant, nor have we included measures of cellular immunity. These analyses are on-going.

In conclusion, we report that both age and frailty impair antibody responses to the primary series of SARS-CoV-2 vaccination in older males, and that these disparities are largely eliminated by receipt of a third vaccine dose. Given that male sex is an important risk factor for severe outcomes from COVID-19 [10-14], the finding that older and frail males may be vulnerable to breakthrough infections due to low antibody responses, particularly before a third vaccine dose is administered, is of considerable public health importance. These findings emphasize that increasing third dose coverage among older males is crucial to protecting this vulnerable population from SARS-CoV-2.

## Data Availability

All data produced in the present study are available upon reasonable request to the authors

## Acknowledgements

The authors thank the participants, as well as Denise C. Kelly and Eileen Sheridan-Malone for collection of samples. The authors thank Emily Egbert for coordination of samples and healthcare worker patients, and Joseph Shiloach, David Quan, and Yesh Muralidharan for technical assistance.

## References

1. Kang S-J, Jung SI. Age-related morbidity and mortality among patients with COVID-19. Infection & chemotherapy 2020; 52(2): 154.

2. O’Driscoll M, Dos Santos GR, Wang L, et al. Age-specific mortality and immunity patterns of SARS-CoV-2. Nature 2021; 590(7844): 140–5.

3. Chen Y, Klein SL, Garibaldi BT, et al. Aging in COVID-19: Vulnerability, immunity and intervention. Ageing Res Rev 2020: 101205.

4. Polack FP, Thomas SJ, Kitchin N, et al. Safety and efficacy of the BNT162b2 mRNA covid-19 vaccine. New England Journal of Medicine 2020.

5. Baden LR, El Sahly HM, Essink B, et al. Efficacy and Safety of the mRNA-1273 SARS-CoV-2 Vaccine. New England Journal of Medicine 2020.

6. Crooke SN, Ovsyannikova IG, Poland GA, Kennedy RB. Immunosenescence: A systems-level overview of immune cell biology and strategies for improving vaccine responses. Exp Gerontol 2019; 124: 110632.

7. Abe KT, Hu Q, Mozafarihashjin M, et al. Neutralizing antibody responses to SARS-CoV-2 variants in vaccinated Ontario long-term care home residents and workers. medRxiv 2021: 2021.08.06.21261721.

8. Canaday DH, Carias L, Oyebanji O, et al. Reduced BNT162b2 mRNA vaccine response in SARS-CoV-2-naive nursing home residents. medRxiv 2021.

9. Causa R, Almagro-Nievas D, Rivera-Izquierdo M, et al. Antibody Response 3 Months after 2 Doses of BNT162b2 mRNA COVID-19 Vaccine in Residents of Long-Term Care Facilities. Gerontology 2021: 1–7.

10. Salje H, Kiem CT, Lefrancq N, et al. Estimating the burden of SARS-CoV-2 in France. Science 2020; 369(6500): 208–11.

11. Richardson S, Hirsch JS, Narasimhan M, et al. Presenting Characteristics, Comorbidities, and Outcomes Among 5700 Patients Hospitalized With COVID-19 in the New York City Area. JAMA 2020; 323(20): 2052–9.

12. Meng Y, Wu P, Lu W, et al. Sex-specific clinical characteristics and prognosis of coronavirus disease-19 infection in Wuhan, China: A retrospective study of 168 severe patients. PLoS pathogens 2020; 16(4): e1008520.

13. Bauer P, Brugger J, Koenig F, Posch M. An international comparison of age and sex dependency of COVID-19 Deaths in 2020-a descriptive analysis. medRxiv 2021.

14. Scully EP, Haverfield J, Ursin RL, Tannenbaum C, Klein SL. Considering how biological sex impacts immune responses and COVID-19 outcomes. Nat Rev Immunol 2020: 1–6.

15. Gubbels Bupp MR, Potluri T, Fink AL, Klein SL. The Confluence of Sex Hormones and Aging on Immunity. Front Immunol 2018; 9: 1269.

16. Márquez EJ, Chung C-h, Marches R, et al. Sexual-dimorphism in human immune system aging. Nature communications 2020; 11(1): 1–17.

17. Shapiro JR, Li H, Morgan R, et al. Sex-specific effects of aging on humoral immune responses to repeated influenza vaccination in older adults. Npj Vaccines 2021; 6(1): 147.

18. Bischof E, Wolfe J, Klein SL. Clinical trials for COVID-19 should include sex as a variable. The Journal of Clinical Investigation 2020; 130(7).

19. Shapiro JR, Morgan R, Leng SX, Klein SL. Roadmap for sex-responsive influenza and COVID-19 vaccine research in older adults. Frontiers in Aging 2022; in press.

20. Fried LP, Tangen CM, Walston J, et al. Frailty in Older Adults: Evidence for a Phenotype. Journals Gerontology Ser Biological Sci Medical Sci 2001; 56(3): M146–M57.

21. Zhong D, Xiao S, Debes AK, et al. Durability of Antibody Levels After Vaccination With mRNA SARS-CoV-2 Vaccine in Individuals With or Without Prior Infection. JAMA 2021.

22. Klein SL, Pekosz A, Park H-S, et al. Sex, age, and hospitalization drive antibody responses in a COVID-19 convalescent plasma donor population. The Journal of Clinical Investigation 2020.

23. NCI Serological Sciences Network for COVID-19 (SeroNet). Available at: https://www.cancer.gov/research/key-initiatives/covid-19/coronavirus-research-initiatives/serological-sciences-network.

24. Karaba AH, Zhu X, Liang T, et al. A Third Dose of SARS-CoV-2 Vaccine Increases Neutralizing Antibodies Against Variants of Concern in Solid Organ Transplant Recipients. American Journal of Transplantation 2021.

25. Newson RB. Sensible parameters for univariate and multivariate splines. The Stata Journal 2012; 12(3): 479–504.

26. Kohler H. Novel vaccine concept based on back-boost effect in viral infection. Vaccine 2015; 33(29): 3274–5.

27. Anderson EM, Eiloa T, Goodwin E, et al. SARS-CoV-2 infections elicit higher levels of original antigenic sin antibodies compared to SARS-CoV-2 mRNA vaccinations.

28. Grobben M, Straten Kvd, Brouwer PJM, et al. Cross-reactive antibodies after SARS-CoV-2 infection and vaccination. eLife 2021; 10: e70330.

29. Bates TA, Leier HC, Lyski ZL, et al. Age-Dependent Neutralization of SARS-CoV-2 and P.1 Variant by Vaccine Immune Serum Samples. JAMA 2021; 326(9).

30. Jabal KA, Ben-Amram H, Beiruti K, et al. Impact of age, ethnicity, sex and prior infection status on immunogenicity following a single dose of the BNT162b2 mRNA COVID-19 vaccine: real-world evidence from healthcare workers, Israel, December 2020 to January 2021. Eurosurveillance 2021; 26(6): 2100096.

31. Collier DA, Ferreira IATM, Kotagiri P, et al. Age-related immune response heterogeneity to SARS-CoV-2 vaccine BNT162b2. Nature 2021; 596(7872): 417–22.

32. Wang J, Tong Y, Li D, Li J, Li Y. The Impact of Age Difference on the Efficacy and Safety of COVID-19 Vaccines: A Systematic Review and Meta-Analysis. Front Immunol 2021; 12.

33. Mwimanzi F, Lapointe H, Cheung PK, et al. Older Adults Mount Less Durable Humoral Responses to a Two-dose COVID-19 mRNA Vaccine Regimen, but Strong Initial Responses to a Third Dose. medRxiv 2022.

34. Ríos SS, Romero MM, Zamora EBC, et al. Immunogenicity of the BNT162b2 vaccine in frail or disabled nursing home residents: COVID-A study. J Am Geriatr Soc 2021; 69(6): 1441–7.

35. Seiffert P, Konka A, Kasperczyk J, et al. Immunogenicity of the BNT162b2 mRNA COVID-19 vaccine in older residents of a long-term care facility: relation with age, frailty and prior infection status. Biogerontology 2021.

36. Demaret J, Corroyer-Simovic B, Alidjinou EK, et al. Impaired Functional T-Cell Response to SARS-CoV-2 After Two Doses of BNT162b2 mRNA Vaccine in Older People. Front Immunol 2021; 12(4639).

37. Hollinghurst J, North L, Perry M, et al. COVID-19 infection risk amongst 14,104 vaccinated care home residents: a national observational longitudinal cohort study in Wales, UK, December 2020–March 2021. Age Ageing 2021.

38. Antonelli M, Penfold RS, Merino J, et al. Risk factors and disease profile of post-vaccination SARS-CoV-2 infection in UK users of the COVID Symptom Study app: a prospective, community-based, nested, case-control study. Lancet Infect Dis 2021.

39. Tan CW, Chia WN, Qin X, et al. A SARS-CoV-2 surrogate virus neutralization test based on antibody-mediated blockage of ACE2–spike protein–protein interaction. Nat Biotechnol 2020; 38(9): 1073–8.

40. Goldblatt D, Fiore-Gartland A, Johnson M, et al. Towards a population-based threshold of protection for COVID-19 vaccines. Vaccine 2021.

41. Carreño JM, Alshammary H, Tcheou J, et al. Activity of convalescent and vaccine serum against SARS-CoV-2 Omicron. Nature 2021.

42. Bartsch Y, Tong X, Kang J, et al. Preserved Omicron Spike specific antibody binding and Fc-recognition across COVID-19 vaccine platforms. medRxiv 2021.

43. Chan CEZ, Seah SGK, Chye DH, et al. The Fc-mediated effector functions of a potent SARS-CoV-2 neutralizing antibody, SC31, isolated from an early convalescent COVID-19 patient, are essential for the optimal therapeutic efficacy of the antibody. Plos One 2021; 16(6): e0253487.

44. Gorman MJ, Patel N, Guebre-Xabier M, et al. Fab and Fc contribute to maximal protection against SARS-CoV-2 following NVX-CoV2373 subunit vaccine with Matrix-M vaccination. Cell Reports Medicine 2021; 2(9).

45. Vorland CJ. Statistics: Sex difference analyses under scrutiny. Elife 2021; 10: e74135.

46. Kuo H, Shapiro JR, Dhakal S, et al. Sex-specific effects of age and body mass index on antibody responses to seasonal influenza vaccines in healthcare workers. Vaccine 2021.

47. Shattuck-Heidorn H, Danielsen AC, Gompers A, et al. A finding of sex similarities rather than differences in COVID-19 outcomes. Nature 2021; 597(7877): E7–E9.

48. Shapiro JR, Klein SL, Morgan R. Stop ‘controlling’ for sex and gender in global health research. Bmj Global Heal 2021; 6(4): e005714.

49. Shapiro JR, Klein SL, Morgan R. COVID-19: use intersectional analyses to close gaps in outcomes and vaccination. Nature 2021; 591(7849): 202.

