## Supplemental for "Association of frailty, age, and biological sex with SARS-CoV-2 mRNA vaccine-induced immunity in older adults"

### Supplemental Materials

Supplemental Methods

**SARS-CoV-2 ELISAs**

The ELISA protocol was adapted from a protocol published by the Florian Krammer laboratory [1]. Antigens were either enigineered at Johns Hopkins as previously described [2] or were obtained through the NCI Serological Sciences Network for COVID-19 [3] (**Supplemental Table 1**). S and S-RBD antigens were diluted to 2ug/ml in PBS. Nucleocapsid antigen was diluted to 1ug/ml in PBS. Ninety-six-well plates (Immulon 4HBX, Thermo Fisher Scientific) were coated overnight at 4°C with 50µL of the antigen solution. Plates were then washed. All washing steps consisted of 3 washes with 300µL PBS plus 0.1% Tween-20 (PBST) (Thermo Fisher Scientific). Plates were blocked with 200µL PBST with 3% nonfat milk (milk powder, American Bio) for 1 hour at room temperature (RT). Plasma samples were heat inactivated at 56°C for 1 hour and then diluted to 1:20 in PBST + 1% nonfat milk before performing ten 3-fold serial dilutions. Pre-pandemic plasma samples were diluted to 1:100 and used as negative controls. A monoclonal antibody against the SARS–CoV-2 S protein (1:5000; catalog 40150-D001, Sino Biological) was used as a positive control for vaccine, Alpha and Delta S and S-RBD ELISAs. Convalescent plasma diluted to 1:100 was used as a positive control for N and Omicron ELISAs. 100µL of samples and controls were added to ELISA plates in duplicate and incubated for 2 hours at RT. Plates were then washed and 50µL of Fc-specific total IgG HRP (1:5000 dilution in PBST plus 1% nonfat milk, catalog A18823, Invitrogen, Thermo Fisher Scientific) was added and incubated at RT for 1 hour. Plates were washed and 100µL of SIGMAFAST OPD (*o*-phenylenediamine dihydrochloride) solution (MilliporeSigma) was added to each well and incubated in the dark at RT for 10 minutes. To stop the reaction, 50µL of 3M HCl (Thermo Fisher Scientific) was added to each well. The OD of each plate was read at 490 nm ($OD_{490}$) on a SpectraMax i3 ELISA Plate Reader (BioTek Instruments). The cutoff value for each plate was calculated by adding the average of negative control OD values to three times the standard deviation of the negative control OD values. For each sample, the highest dilution above the cut-off value was considered the endpoint titer. The cut-off value was then subtracted from all sample OD values, and negative values set to zero. Background-subtracted OD values were plotted against the dilution factor to calculate the area under the curve (AUC). For each S and S-RBD antigen, the limit of detection was set to half of the lowest measured AUC for samples with a detectable titer (i.e., titer ≥ 20). For samples with an undetectable titer (i.e., titer < 20), the AUC was arbitrarily set to half of the limit of detection. For nucleocapsid assays, the threshold for seropositivity was set to a titer of 1:180 based on pre-pandemic samples.

**ACE2 inhibition Assays**

The MSD ACE2 inhibition assay measures the ability of participant plasma to inhibit ACE2 binding to spike protein. Plasma was thawed and ACE2 blocking was measured using the ACE2 MSD V-PLEX SARS-CoV-2 ACE2 kits according to the manufacturer's protocol at a dilution of 1:100. Plates come pre-coated with spike proteins corresponding to variants of interest. They were washed and incubated with plasma for one hour, human ACE2 protein conjugated with a SULFO-TAG (light-emitting label) added for another hour, washed, read buffer added, and read with a MESO QuickPlex SQ 120 instrument. An 8-point calibration curve was included on each plate such that results could be expressed as ACE2-inhibiting activity corresponding to 1 µg/ml of monoclonal antibody to SARS-CoV-2 Spike protein. Values below the manufacturer-specified limit of detection of 0.448 µg/ml were arbitrarily set to half of that value.

Supplemental tables

**Supplemental table 1. SARS-CoV-2 antigens for ELISAs**

| Antigen | Protein name [relevant mutations] | Source |
|---|---|---|
| Nucleocapsid | SARS-CoV-2 Nucleocapsid (N) C-terminal domain (247-364) | SeroNet |
| Vaccine S | ECD ΔFurin HexaPro [4] | JHU |
| Vaccine RBD | As previously described [1] | JHU |
| Alpha S | hCoV19/USA/MD-HP01101/2021 (EPI_ISL_825013) [H69del, V70del, Y145del, N501Y, A570D, D614G, P681H, T716I, S982A, D1118H] | JHU |
| | SARS-CoV-2 B.1.1.7 S-2P(15-1208)-T4f-3C-His8-Strep2x2 [Δ(69-70), Δ144, N501Y, A570D, D614G, P681H, T716I, S982A, D1118H] | SeroNet |
| Delta S | hCoV-19/USA/MD-HP05285/2021 (EPI_ISL_2103264; strain B.1.617.2a) [T19R, G142D, E156G, F157del, R158del, A222V, L452R, T478K, D614G, P681R, D950N] | JHU |
| | SARS-CoV-2 B.1.617.2 S-2P(15-1208)-T4f-3C-His8-Strep2x2 [T19R, Δ(157-158), L452R, T478K, D614G, P681R, D950N] | SeroNet |
| Omicron S | SARS-CoV-2-S(1-1208)-2P-3C-His8-Strep2x2 B.1.1.529 [A67V, DEL(69-70), T95I, G142D, Δ(143-145), Δ 211, L212I, ins(214)-EPE, G339D, S371L, S373P, S375F, K417N, N440K, G446S, S477N, T478K, E484A, Q493R, G496S, Q498R, N501Y, Y505H, T547K, D614G, H655Y, N679K, P681H, N764K, D796Y, N856K, Q954H, N969K, L981F] | SeroNet |

**Supplemental table 2. Anti-vaccine strain IgG geometric mean titers in older adults**

| GMT (95% CI) | All | Males | Females |
|---|---|---|---|
| **Vaccine S IgG** | | | |
| Pre | 39 (31-49) | 43 (30-62) | 36 (26-49) |
| <1M_PD1 | 3827 (1481-9894) | 2186 (463-10313) | 6396 (1740-23508) |
| <1M_PD2 | 67232 (47320-95524) | 45490 (25336-81677) | 87790 (56712-135897) |
| 3M_PD2 | 17354 (12375-24336) | 10219 (5956-17532) | 25253 (16656-38288) |
| 6M_PD2 | 5575 (4303-7225) | 4115 (2750-6157) | 6901 (4927-9667) |
| 1M_PD3 | 131220 (101585-169501) | 120586 (81207-179062) | 139980 (98411-199108) |
| **Vaccine S-RBD IgG** | | | |
| Pre | 34 (28-40) | 40 (31-53) | 30 (24-37) |
| <1M_PD1 | 871 (357-2126) | 489 (160-1493) | 1478 (339-6442) |
| <1M_PD2 | 16299 (10293-25810) | 11983 (5503-26094) | 20110 (11242-35971) |
| 3M_PD2 | 3041 (2175-4251) | 2311 (1247-4283) | 3693 (2523-5405) |
| 6M_PD2 | 1512 (1091-2096) | 1123 (679-1857) | 1864 (1208-2876) |
| 1M_PD3 | 57565 (41091-80643) | 45628 (25164-82732) | 68761 (46015-102751) |
| **Vaccine ACE2-inhibition** | | | |
| Pre | 0.2 (0.2-0.2) | 0.2 (0.2-0.2) | 0.2 (0.2-0.2) |
| <1M_PD1 | 2.3 (1.4-3.7) | 1.6 (0.7-3.6) | 3.1 (1.7-5.8) |
| <1M_PD2 | 13.8 (9.5-19.9) | 11.1 (6.1-20.3) | 16.0 (9.9-25.9) |
| 3M_PD2 | 5.6 (4.2-7.4) | 3.9 (2.3-6.5) | 7.1 (5.2-9.7) |
| 6M_PD2 | 0.5 (0.4-0.7) | 0.4 (0.3-0.7) | 0.6 (0.3-0.9) |
| 1M_PD3 | 93.5 (60.2-145.1) | 80.5 (43.3-149.8) | 104.7 (55.2-198.8) |

**Supplemental table 3. Anti-Alpha, Delta, and Omicron IgG geometric mean titers in older adults**

| GMT (95% CI) | All | Males | Females |
|---|---|---|---|
| **Alpha S IgG** | | | |
| <1M_PD1 | 1005 (472-2139) | 597 (178-2003) | 1620 (578-4542) |
| <1M_PD2 | 19112 (12834-28461) | 14580 (7894-26928) | 22993 (13438-39340) |
| 3M_PD2 | 7768 (5707-10572) | 5531 (3334-9174) | 9880 (6713-14542) |
| 6M_PD2 | 1894 (1398-2566) | 1913 (1264-2896) | 1880 (1211-2919) |
| 1M_PD3 | 101108 (78206-130716) | 88356 (58551-133334) | 111644 (79308-157164) |
| **Alpha ACE2-inhibition** | | | |
| <1M_PD1 | 1.7 (1.1-2.7) | 1.4 (0.7-2.7) | 2.1 (1.0-4.2) |
| <1M_PD2 | 11.0 (7.8-15.6) | 8.2 (4.4-15.2) | 13.5 (8.9-20.6) |
| 3M_PD2 | 5.5 (4.5-6.8) | 4.2 (2.8-6.3) | 6.6 (5.3-8.4) |
| 6M_PD2 | 0.4 (0.3-0.5) | 0.3 (0.2-0.4) | 0.5 (0.3-0.8) |
| 1M_PD3 | 85.7 (53.2-137.9) | 68.7 (32.2-146.6) | 101.4 (53.6-191.7) |
| **Delta S IgG** | | | |
| <1M_PD1 | 1106 (535-2282) | 729 (211-2515) | 1620 (632-4152) |
| <1M_PD2 | 19112 (12935-28240) | 12961 (6844-24546) | 24917 (15155-40968) |
| 3M_PD2 | 7768 (5694-10596) | 5184 (3093-8689) | 10343 (7081-15108) |
| 6M_PD2 | 1808 (1394-2345) | 1418 (952-2113) | 2145 (1517-3031) |
| 1M_PD3 | 80862 (61259-106738) | 77443 (50049-119831) | 83472 (57148-121920) |
| **Delta ACE2-inhibition** | | | |
| <1M_PD1 | 1.1 (0.5-2.3) | 0.8 (0.3-2.1) | 1.5 (0.5-5.1) |
| <1M_PD2 | 11.1 (7.2-17.2) | 7.1 (3.3-15.5) | 15.2 (9.1-25.4) |
| 3M_PD2 | 6.4 (4.8-8.6) | 4.9 (3.0-8.0) | 7.7 (5.4-11.0) |
| 6M_PD2 | 0.4 (0.3-0.5) | 0.3 (0.2-0.4) | 0.4 (0.3-0.6) |
| 1M_PD3 | 77.0 (49.5-119.9) | 62.9 (30.6-129.2) | 90.0 (50.3-161.0) |
| **Omicron S IgG** | | | |
| <1M_PD1 | 510 (126-2057) | 297 (47-1883) | 1073 (80-14373) |
| <1M_PD2 | 4938 (3386-7201) | 3551 (1947-6475) | 6185 (3776-10132) |
| 3M_PD2 | 2118 (1551-2893) | 1568 (948-2595) | 2620 (1756-3907) |
| 6M_PD2 | 1084 (759-1549) | 699 (372-1311) | 1475 (976-2231) |
| 1M_PD3 | 35761 (26798-47722) | 31194 (19580-49697) | 39699 (27100-58155) |

Supplemental figures

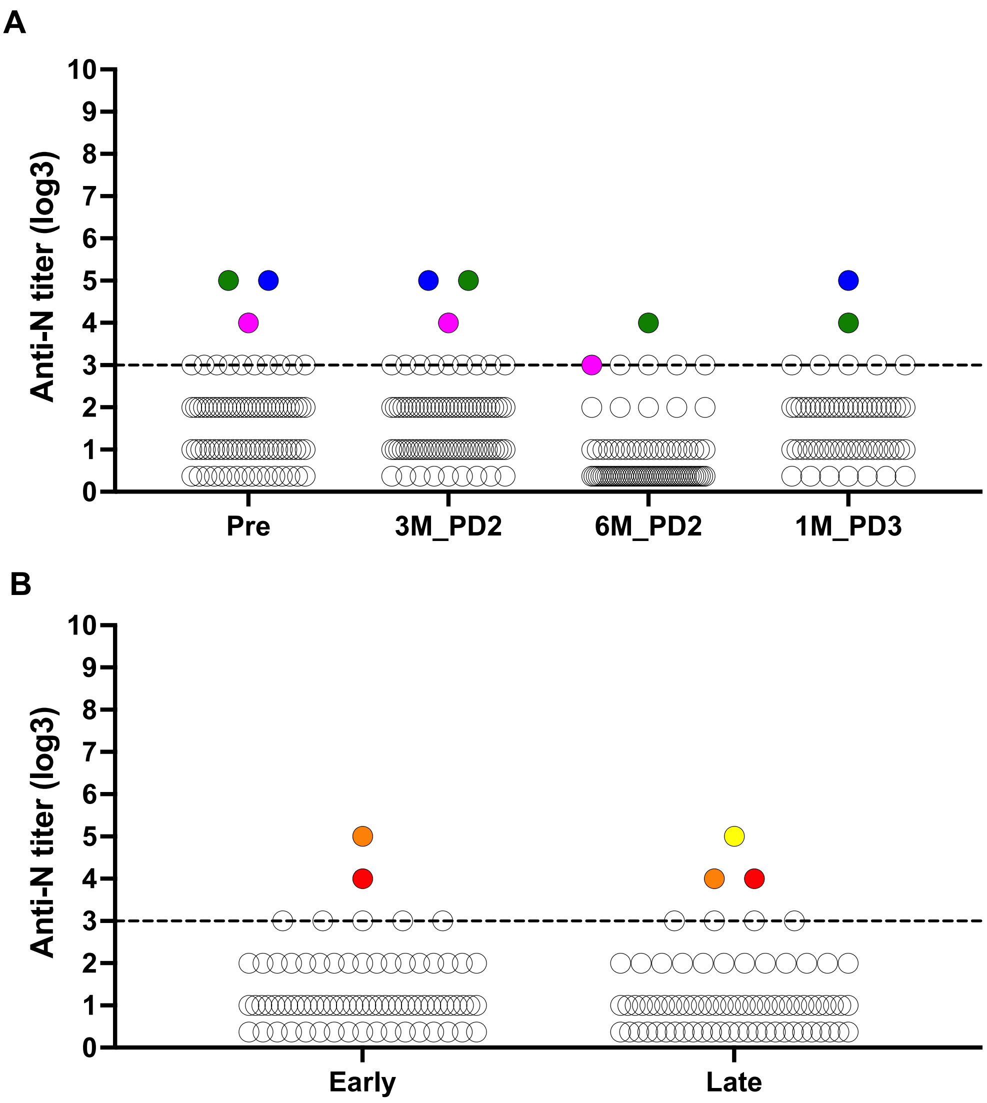

**Supplemental figure 1. Anti-nucleocapsid IgG titers in older and younger adults.**
Anti-nucleocapsid (N) IgG endpoint titers are plotted on the $\log_3$-scale for the older (**A**) and younger (**B**) adults. Dashed lines indicated the threshold for positivity (titer of 1:180), which was established using pre-pandemic samples. Colored dots indicate individuals who were excluded from further analysis due to positive anti-N IgG indicating previous infection, and dots of the same color are data from the same individual over time.

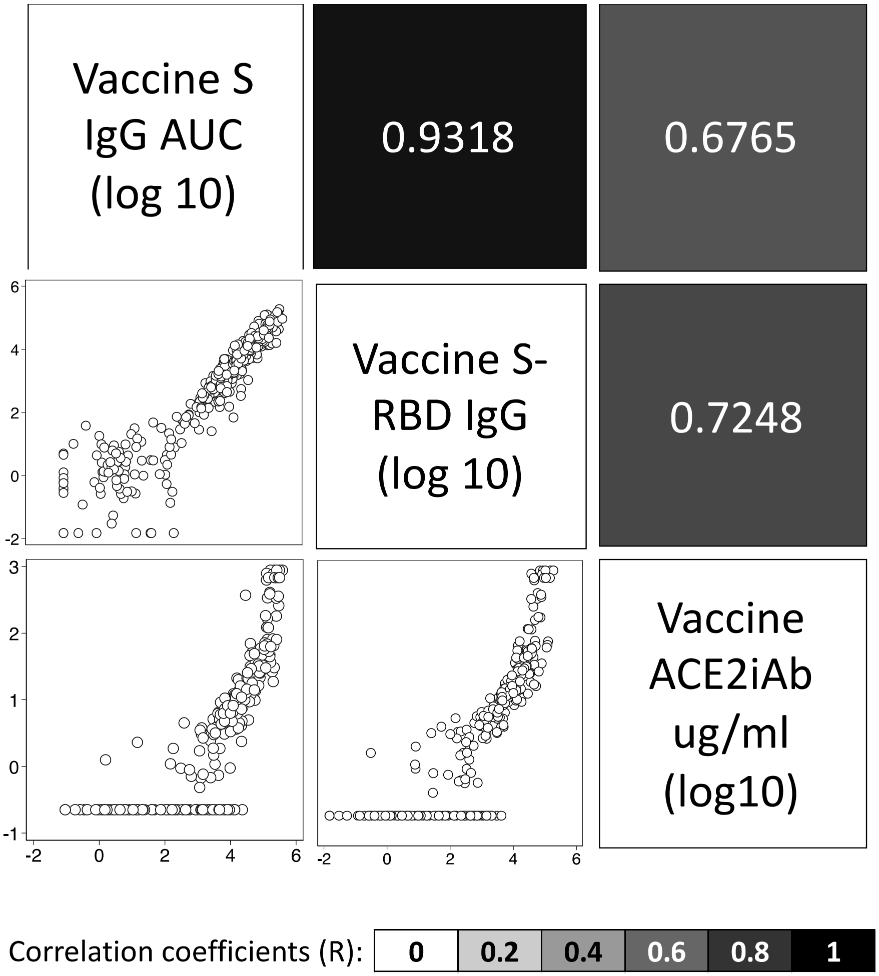

**Supplemental figure 2. Measures of vaccine-induced humoral immunity to the vaccine strain of SARS-CoV-2 are highly correlated with each other in older adults.**

The correlation between anti-S IgG, anti-S-RBD IgG, and ACE2-inhbiting antibodies (ACE2iAb), in older adults was assessed for all study timepoints together. Scatter plots are shown in the lower half of the matrix, and correlation coefficients (R), color coded by the strength of the correlation, are shown in the upper half of the matrix.

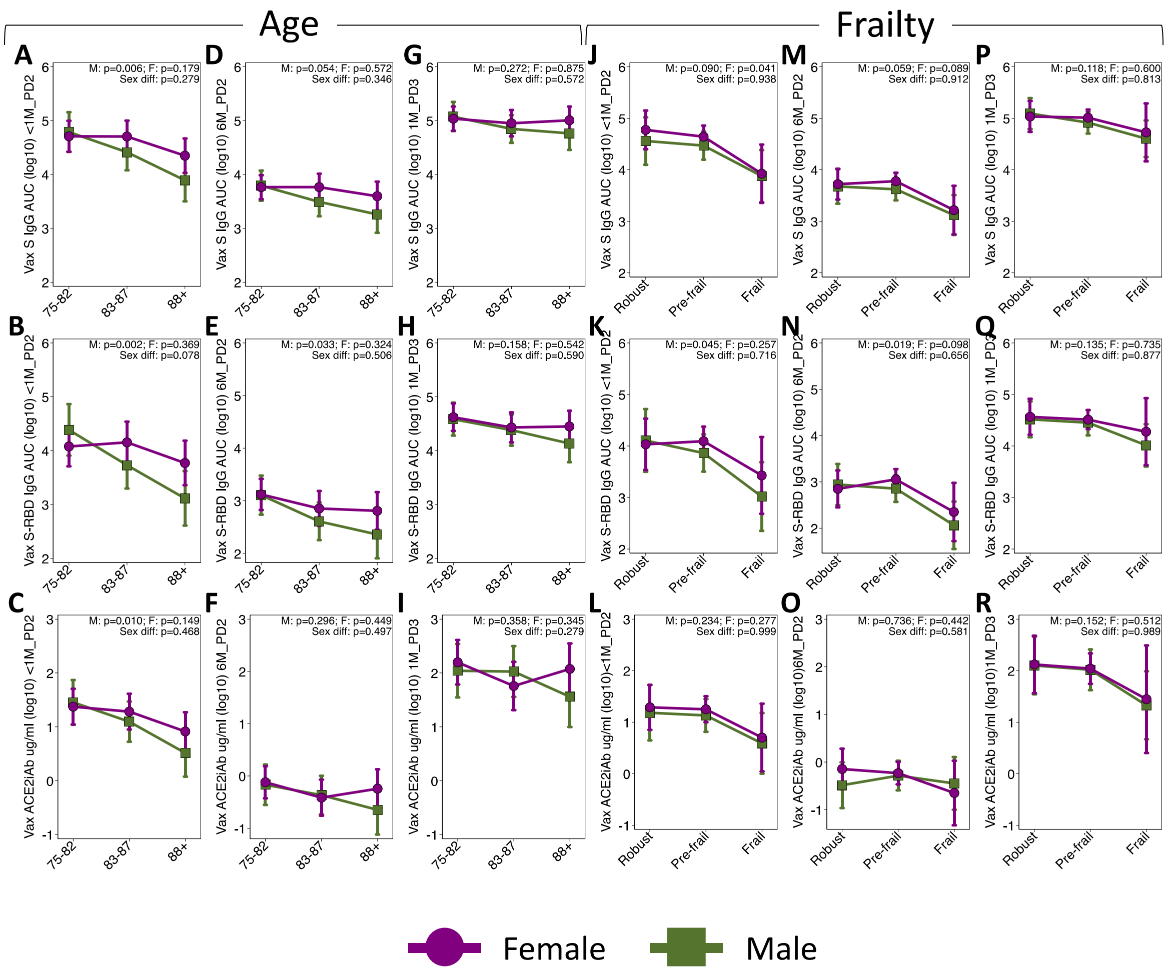

**Supplemental figure 3. Sex-specific effects of aging and frailty 14-30-days and 6-months post dose 2, and 14-30-days post dose 3 in older adults.**

The effect of age on anti-S IgG, anti-S-RBD IgG, and ACE2iAb are shown <1M_PD2 for females (n=15 aged 75-82; n=14 aged 83-87; n=12 aged ≥88) and males (n=9 aged 75-82; n=11 aged 83-87; n=8 aged ≥88) (**A-C**), 6M_PD2 for females (n=19 aged 75-82; n=15 aged 83-87; n=13 aged ≥88) and males (n=12 aged 75-82; n=13 aged 83-87; n=8 aged ≥88) (**D-F**), and 1M_PD3 for females (n=13 aged 75-82; n=11 aged 83-87; n=10 aged ≥88) and males (n=9 aged 75-82; n=10 aged 83-87; n=7 aged ≥88)(**G-I**). Similarly, the effect of frailty on the three measures of humoral immunity are shown for <1M_PD2 for females (n=9 robust; n=28 pre-frail; n=4 frail) and males (n=6 robust; n=17 pre-frail; n=5 frail) (**J-L**), 6M_PD2 for females (n=10 robust; n=33 pre-frail; n=4 frail) and males (n=8 robust; n=19 pre-frail; n=6 frail)(**M-O**), and 1M_PD3 for females (n=7 robust; n=25 pre-frail; n=2 frail) and males (n=7 robust; n=14 pre-frail; n=5 frail) (**P-R**). The effects of age (**A-I**) or frailty (**J-R**) in males and females, and sex-differences in these effects, were tested using linear regression models with interaction terms between sex and age or frailty, and all p-values are shown. All point estimates are accompanied by error bars indicating the 95% confidence interval.

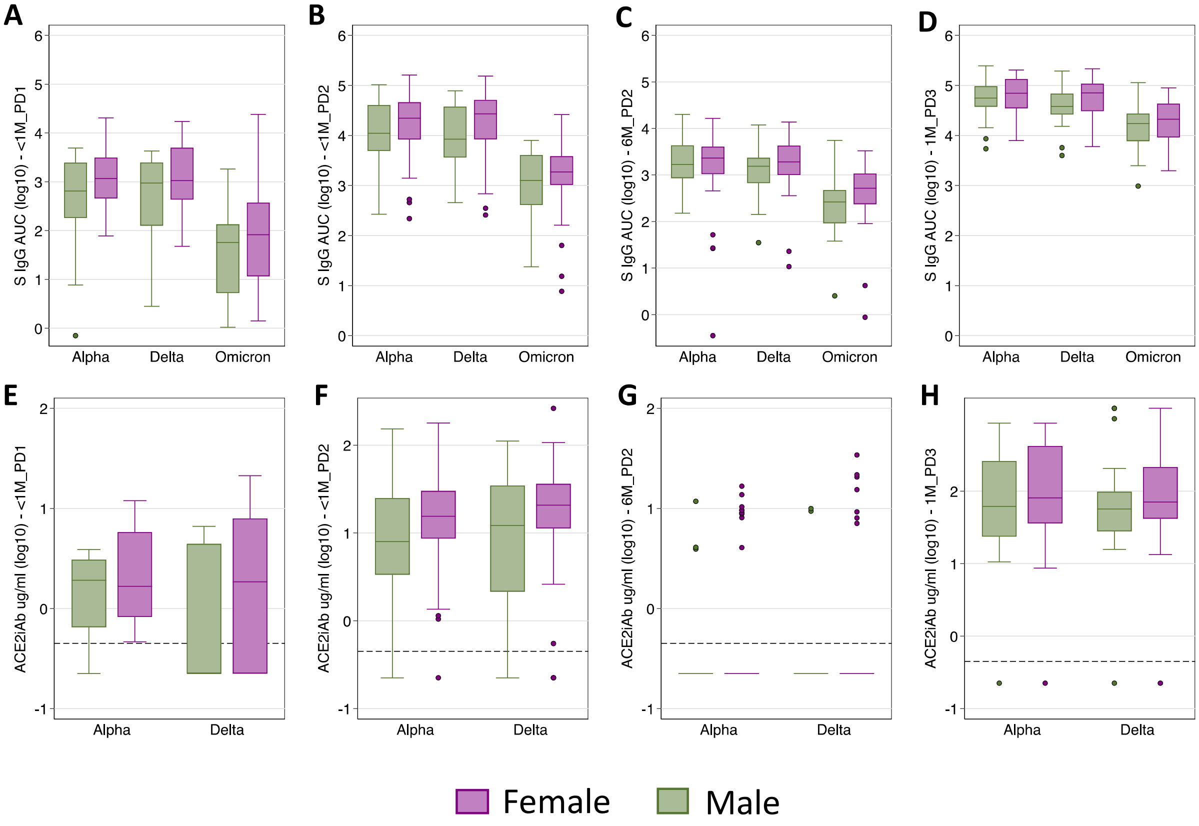

**Supplemental figure 4. Antibody responses to variants of concern tend to be higher in older females than older males.**

Anti-S (**A-D**) and ACE2-inhibiting (**E-H**) antibodies against the Alpha and Delta variants of concern are shown for males and females <1M_PD1 (**A, E**), <1M-PD2(**B, F**), 6M_PD2 (**C, G**), and 1M_PD3 (**D, H**). Dashed lines show the lower limit of detection of the assay.

Supplemental references

1. Stadlbauer D, Amanat F, Chromikova V, et al. SARS-CoV-2 seroconversion in humans: a detailed protocol for a serological assay, antigen production, and test setup. Current protocols in microbiology **2020**; 57(1): e100.
2. Klein SL, Pekosz A, Park H-S, et al. Sex, age, and hospitalization drive antibody responses in a COVID-19 convalescent plasma donor population. The Journal of Clinical Investigation **2020**.
3. NCI Serological Sciences Network for COVID-19 (SeroNet). Available at: https://www.cancer.gov/research/key-initiatives/covid-19/coronavirus-research-initiatives/serological-sciences-network.
4. Hsieh C-L, Goldsmith JA, Schaub JM, et al. Structure-based design of prefusion-stabilized SARS-CoV-2 spikes. Science **2020**; 369(6510): 1501-5.
